# Genetic and Cellular Architecture of Breast Cancer Risk Across Ancestries

**DOI:** 10.1101/2025.08.20.25334075

**Authors:** James L. Li, Maria Zanti, Jacob Williams, Om Jahagirdar, Guochong Jia, Alistair Turcan, Qiang Hu, Jean-Tristan Brandenburg, Li Yan, Weang-Kee Ho, Jingmei Li, José Patricio Miranda, Devika Godbole, Julie-Alexia Dias, Xiaomeng Zhang, Leila Dorling, Wenlong Carl Chen, Nicholas Boddicker, Ying Wang, Alicia Martin, Yan Dora Zhang, Joe Dennis, Esther M. John, Gabriela Torres-Mejia, Lawrence H. Kushi, Jeffrey Weitzel, Susan L. Neuhausen, Luis Carvajal-Carmona, Christopher Haiman, Elad Ziv, Laura Fejerman, Wei Zheng, Dezheng Huo, Douglas Easton, Stephen J. Chanock, Nilanjan Chatterjee, Peter Kraft, Montserrat Garcia-Closas, Wendy S.W. Wong, Kyriaki Michailidou, Qianqian Zhu, Martin Jinye Zhang, Diptavo Dutta, Thomas U. Ahearn, Haoyu Zhang

**Affiliations:** Department of Public Health Sciences, University of Chicago, Chicago, IL, USA; Department of Biostatistics, The Cyprus Institute of Neurology and Genetics, Nicosia, Cyprus; Division of Cancer Epidemiology and Genetics, National Cancer Institute, National Institutes of Health, Bethesda, MD, USA; Division of Epidemiology, Department of Medicine, Vanderbilt Epidemiology Center, Vanderbilt-Ingram Cancer Center, Vanderbilt University Medical Center, Nashville, TN, USA; Ray and Stephanie Lane Computational Biology Department, School of Computer Science, Carnegie Mellon University, Pittsburgh, PA, USA; Department of Biostatistics and Bioinformatics, Roswell Park Comprehensive Cancer Center, Buffalo, NY, USA; Sydney Brenner Institute for Molecular Bioscience, Faculty of Health Sciences, University of the Witwatersrand, Johannesburg, South Africa; Strengthening Oncology Services Research Unit, Faculty of Health Sciences, University of the Witwatersrand, Johannesburg, South Africa; School of Mathematical Sciences, Faculty of Science and Engineering, University of Nottingham Malaysia, Semenyih, Selangor, Malaysia; Cancer Research Malaysia, Subang Jaya, Selangor, Malaysia; Genome Institute of Singapore, Agency for Science, Technology and Research (A*STAR), Singapore; Department of Nutrition, Diabetes and Metabolism, School of Medicine, Pontificia Universidad Católica de Chile, Santiago, Chile; PhD Program in Epidemiology, Pontificia Universidad Católica de Chile, Santiago, Chile; Advanced Center for Chronic Diseases, Pontificia Universidad Católica de Chile & Universidad de Chile, Santiago, Chile; Department of Biostatistics, Harvard T.H. Chan School of Public Health, Boston, MA, USA; Centre for Cancer Genetic Epidemiology, Department of Public Health and Primary Care, University of Cambridge, Cambridge, UK; Department of Quantitative Health Sciences, Mayo Clinic, Rochester, MN, USA; Analytic and Translational Genetics Unit, Massachusetts General Hospital, Boston, MA, USA; Department of Medicine, Harvard Medical School, Boston, MA, USA; Stanley Center for Psychiatric Research, Broad Institute of MIT and Harvard, Cambridge, MA, USA; Department of Statistics & Actuarial Science, School of Computing and Data Science, The University of Hong Kong, Hong Kong SAR, China; Departments of Epidemiology & Population Health and of Medicine (Oncology), Stanford University School of Medicine, Stanford, CA, USA; Stanford Cancer Institute, Stanford University School of Medicine, Stanford, CA, USA; Instituto Nacional de Salud Pública, Cuernavaca, Mexico; UC Davis Genome Center, University of California, Davis, Davis, CA, USA; Division of Research, Kaiser Permanente Northern California, Oakland, CA, USA; Division of Precision Prevention, University of Kansas Comprehensive Cancer Center, Kansas City, KS, USA; Department of Population Sciences, Beckman Research Institute of City of Hope, Duarte, CA, USA; Department of Biochemistry and Molecular Medicine, School of Medicine, University of California at Davis, Davis, CA, USA; The Health Equity, Leadership, Science and Community Laboratory, Genome Center, University of California at Davis, Davis, CA, USA; Department of Preventive Medicine, Norris Comprehensive Cancer Center, Keck School of Medicine, University of Southern California, Los Angeles, CA, USA; Division of General Internal Medicine, Department of Medicine, University of California, San Francisco, San Francisco, CA, USA; Department of Public Health Sciences, University of California Davis, Davis, CA, USA; Genome Center, University of California Davis, Davis, CA, USA; UC Davis Comprehensive Cancer Center, University of California Davis, Davis, CA, USA; Centre for Cancer Genetic Epidemiology, Department of Oncology, University of Cambridge, Cambridge, UK; Department of Biostatistics, Johns Hopkins University, Baltimore, MD, USA; Department of Oncology, Johns Hopkins School of Medicine, Baltimore, MD, USA; Cancer Epidemiology and Prevention Research Unit, The Institute of Cancer Research and Imperial College London, London, UK; National Cancer Registry, a Division of the National Institute for Communicable Diseases, National Health Laboratory Service, Johannesburg, South Africa

**Keywords:** Breast cancer, GWAS, multi-ancestry, SNP-based heritability, Genetic correlation, Single-cell RNA-seq

## Abstract

**Background:** Breast cancer genome-wide association studies (GWAS) have identified more than 200 susceptibility loci, but most studies are dominated by European and East Asian populations.

**Methods:** We analyzed breast cancer GWAS summary statistics from African (AFR), East Asian (EAS), European (EUR), and Hispanic/Latina (H/L) samples (159,297 cases and 212,102 controls). We estimated logit-scale SNP-based heritability, polygenicity, and cross-ancestry genetic correlation, partitioned heritability across functional annotations, and integrated GWAS results with the Tabula Sapiens single-cell atlas using scDRS+.

**Results:** The logit-scale heritability of breast cancer ranged from *h*^2^ =0.47 (SE = 0.07) in EAS to AFR *h*^2^ =0.61 (SE = 0.10), with no significant differences across ancestries (p=0.63). The model-implied number of non-null susceptibility SNPs in the sparse normal-mixture effect-size model also varied from 4,446 (SE = 3,100) in EAS to 8,308 (SE = 2,751) in AFR, but differences were not significant across ancestries (p=0.55). Cross-sample genetic correlations varied, with the strongest correlation between EUR and EAS (*ρ* = 0.79, SE = 0.08) and weakest between AFR and H/L (*ρ* = 0.26, SE = 0.24). Regulatory annotations were enriched for breast cancer heritability across samples. Integration with single-cell expression profiles implicated ancestry-shared associations with innate immune, secretory epithelial, and stromal cell types.

**Conclusion:** These results indicate substantial cross-ancestry sharing of breast cancer polygenic architecture, highlight a consistent contribution of regulatory variation, and identify convergent cellular contexts that motivate functional follow-up and inform expectations for the transferability and attainable performance of common-variant risk prediction across populations.

## INTRODUCTION

Breast cancer is the most common cancer among women worldwide, affecting over two million individuals annually^1^. Genome-wide association studies (GWAS) have identified more than 200 genome-wide significant susceptibility markers associated with breast cancer risk^2–7^. However, most GWAS have focused on populations of European (EUR) and East Asian (EAS) ancestry samples^2,4–9^, while studies in African (AFR) ^3,10–13^ and Hispanic/Latina (H/L)^14,15^ samples are limited, and often with smaller sample sizes. Existing studies have both replicated risk markers identified across ancestry populations, as well as identified population-specific risk markers. Furthermore, polygenic risk score (PRS) models trained in European samples often exhibit lower predictive performance in other genetic ancestry samples^16–18^, emphasizing the need for a broader understanding of breast cancer genetics across different ancestries.

Despite this recognition, the reasons why PRS performance varies across ancestries remain incompletely characterized. Potential genetic and technical contributors include differences in GWAS sample size, LD structure, allele-frequency spectra, imputation and variant tagging, heritability, and effect-size distributions. Breast cancer subtype distributions may also matter because genetic associations and PRS performance differ by subtype^4,19,20^. In addition, population differences in lifestyle, anthropometric, and reproductive risk factors can influence absolute-risk estimates and calibration when PRS are incorporated into multifactorial risk models^21^. Harmonized comparisons of genetic architecture are therefore needed both to distinguish these contributions and to project how PRS performance may improve as diverse GWAS resources grow.

We examined the genetic architecture of breast cancer using GWAS summary statistics from 159,297 cases and 212,102 controls from AFR, EAS, EUR, and H/L samples (**Table 1; Figure S1**). We estimated genome-wide SNP-based heritability, characterized the effect-size distribution and model-implied number of non-null susceptibility SNPs, and assessed genetic correlations across samples. We then used GENESIS to project expected CT-PRS performance across hypothetical GWAS sample sizes, translating the estimated architecture into sample-size-dependent prediction trajectories rather than constructing a new PRS. Finally, we examined functional enrichment and used scDRS+ to identify cell types implicated in breast cancer across ancestry samples.

**Table 1.** Sample-specific estimates of the logit-scale SNP-based heritability.

| Sample | Cases | Controls | MAF cutoff | LD Score | No. of SNPs | Heritability<sup>e</sup> |
| --- | --- | --- | --- | --- | --- | --- |
| AFR<sup>a</sup> | 18,034 | 22,104 | MAF > 0.01 | AABCG | 1,628,778 | 0.614 (0.095) |
| EAS<sup>b</sup> | 20,393 | 86,329 | MAF > 0.01 | 1000G | 1,189,628 | 0.466 (0.066) |
| EUR<sup>c</sup> | 118,474 | 96,201 | MAF > 0.01 | 1000G | 1,415,774 | 0.501 (0.050) |
| H/L<sup>d</sup> | 2,396 | 7,468 | MAF > 0.05 | 1000G | 1,138,603 | 0.588 (0.360) |
| Total<sup>f</sup> | 159,297 | 212,102 |  |  |  |  |

Summary-statistic sources: <sup>a</sup> African Ancestry Breast Cancer Genetics (AABCG) consortium; <sup>b</sup> Breast Cancer Association Consortium (BCAC) (OncoArray) and Biobank Japan; <sup>c</sup> BCAC (iCOGs and OncoArray); <sup>d</sup> NC-BCFR/SFBCS (Northern California Breast Cancer Family Registry/ San Francisco Bay Area Breast Cancer Study), RPGEH (Research project on genes environment and health), MEC (Multiethnic cohort), COLUMBUS and CAMA (Cancer de mama) studies/consortia. Variants included in the analyses were on HapMap3 + Multi-Ethnic Genotyping Array (MEGA). <sup>e</sup> logit-scale heritability (standard error). The logit-scale heritability (also known as frailty-scale heritability) is defined as $\sigma_{GWAS}^2 = Var(\sum_{m=1}^M \beta_m G_m)$ , where $G_m$ is the standardized genotype for the mth SNP, $\beta_m$ is the true log odds ratio for the mth SNP and M is the total number of causal SNPs among the GWAS variants. AFR, African; EAS, East Asian; EUR, European; H/L, Hispanic and Latina; MAF, minor allele frequency; LD, linkage disequilibrium; SNPs, single nucleotide polymorphisms. <sup>f</sup> Total case and control counts represent the sums across the ancestry-specific GWAS datasets.

To our knowledge, this study provides: (1) the first harmonized cross-ancestry comparison of logit-scale SNP-based heritability using consistent methodology across AFR, EAS, EUR, and H/L samples; (2) model-based comparisons of polygenicity and projected CT-PRS performance that distinguish the contributions of architecture versus sample size to cross-ancestry prediction gaps; and (3) integration with scDRS+ to identify convergent cellular contexts across ancestries as a biologically interpretable layer of the GWAS signals.

## METHODS

### Study samples and GWAS summary statistics

We analyzed breast cancer GWAS summary statistics from multiple international consortia, totaling 159,297 cases and 212,102 controls across four ancestry groups, African (AFR), East Asian (EAS), European (EUR), and Hispanic/Latina (H/L) (**Table S1**). Sample labels (AFR, EAS, EUR, H/L) were based on published descriptions of the contributing GWAS. AFR data were obtained from African Ancestry Breast Cancer Genetics (AABCG) consortium (18,034 cases and 22,104 controls)^12^. For EAS, we conducted a fixed-effect meta-analysis combining the GWAS summary statistics generated from the Breast Cancer Association Consortium (BCAC)^7^ and Biobank Japan^22^ (20,393 cases and 86,329 controls). EUR summary statistics were obtained from BCAC (118,474 cases and 96,201 controls)^4^. H/L ancestry summary statistics included 2,396 cases and 7,468 controls^15^. Summary-statistic processing and harmonization steps are summarized in **Figure S1**.

### Linkage disequilibrium reference panels and LD score construction

We constructed ancestry-specific linkage-disequilibrium (LD) score reference panels using unrelated individuals from Phase 3 of the 1000 Genomes Project (1000G)^23^, excluding first- and second-degree relatives as well as individuals with first-cousin-level relatedness due to inbreeding^24^. For the EUR, EAS, and H/L samples, LD scores were computed using 484 EUR, 480 EAS, and 317 AMR individuals in the 1000G reference panel, respectively. For the AFR sample, we estimated LD scores using a set of 2,730 AFR women controls from the AABCG consortium. As a sensitivity analysis, we also generated LD scores using 575 AFR individuals from 1000G. LD scores were computed using LD Score Regression (LDSC)^25^ with a LD window of 1MB using the flag “--l2 --ld-window-kb 1000”.

### SNP-based heritability estimation

We estimated logit-scale SNP-based heritability of breast cancer risk within each ancestry using LDSC^25^. On the logit scale, heritability is defined as: σ^2^_*GWAS*_ = *Var*(Σ^*M*^_*m*_=1 β_*m*_*G_m_*), where *G_m_* is the standardized genotype for the *m*th SNP, *β_m_* is the underlying log odds ratio, and M is the total number of causal SNPs among the GWAS variants. This quantity, also referred to as frailty-scale heritability, represents the variance in log-odds of disease explained by common genetic variants. To estimate the logit-scale heritability, we computed the effective sample size as follows:

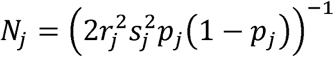

where *s_j_* is the standard error of the log odds ratio, and *p_j_* is the effect allele frequency for variant *j*, and *r*^2^_*j*_ is the imputation quality score for variant *j* (*r*^2^_*j*_ = 1 for directly genotyped variants). To limit the influence of low-information variants, we excluded variants in the lowest 10% of effective sample size within each summary-statistic dataset.

Because the AFR GWAS summary statistics were predominantly derived from U.S.-based studies (85.3%)^12^, we used AABCG controls as the primary AFR LD reference to better match the LD structure and additionally report results using 1000 Genomes AFR as a sensitivity analysis. To assess robustness, we evaluated MAF thresholds of 0.01 and 0.05 in ancestry-specific heritability sensitivity analyses using the HapMap3 ^26^ and HapMap3 plus Multi-Ethnic Genotyping Array (MEGA)^27^ variant sets. Because of the relatively small H/L sample, variants with MAF between 0.01 and 0.05 had lower minor-allele counts and were expected to have less reliable allele-frequency, association-effect, and LD estimates. We therefore used MAF > 0.05 for subsequent H/L analyses to focus inference on more reliably estimated common variants and used HapMap3+MEGA as the primary H/L heritability setting to retain broader common-variant coverage.

### Polygenicity and projected PRS performance

To characterize polygenicity, we applied the GENESIS framework^28,29^, which models GWAS summary statistics using a three-component mixture distribution for SNP effect sizes. Following GENESIS recommendations, we restricted analyses in all ancestry groups to HapMap3 variants with MAF > 0.05. For EAS and EUR, we used the precomputed LD inputs distributed with GENESIS; for AFR and H/L, we constructed ancestry-matched LD inputs using the reference sources described above. From the fitted model, GENESIS estimated the model-implied number of non-null susceptibility SNPs among the analyzed HapMap3 variants. This model-implied count is defined over the analyzed HapMap3 variants and does not represent the number of LD-independent index SNPs or distinct susceptibility loci. GENESIS also projected clumping-and-thresholding PRS (CT-PRS)^30,31^ performance across increasing GWAS sample sizes, including the expected fraction of SNP-based heritability explained by CT-PRS and the corresponding projected area under the receiver operating characteristic curve (AUC). Additional model-fitting details are provided in the **Supplementary Methods**.

Before mixture-model fitting, GWAS summary statistics were processed using the GENESIS preprocessing function with filter = TRUE. Following the recommended GENESIS quality-control procedure, we removed SNPs with effective sample sizes below 0.67 times the 90th percentile of the SNP-level effective-sample-size distribution, excluded variants in the major histocompatibility complex region, and removed SNP-level extreme-effect outliers with Z^2^ > 80 (|Z| > 8.94).

GENESIS projections estimate expected CT-PRS performance conditional on the fitted ancestry-specific common-variant architecture, LD inputs, analyzed variant set, and training sample size; they are not external validations. Observed AUCs additionally depend on the evaluation population, phenotype and subtype composition, genotyping and imputation, covariate adjustment, and score construction. To assess empirical consistency, we projected CT-PRS AUC at the case-control training sample sizes reported in published ancestry-specific PRS studies and compared the projections with published validation AUCs (**Table S5**).

### Cross-ancestry genetic correlation

We estimated cross-ancestry genetic correlation in breast cancer risk using Popcorn^32^, which estimates genetic effect correlations accounting for LD differences across ancestries. Cross-ancestry LD reference panels were built from the same ancestry-specific sources used in heritability analyses. We applied MAF > 0.01 in AFR, EAS, and EUR and MAF > 0.05 in H/L. Because Popcorn estimates each pair using variants that pass filtering in both datasets, correlations involving H/L were restricted to variants with MAF > 0.05 in the H/L sample. Using default settings, we computed pairwise cross-sample genetic correlations for (1) HapMap3 variants and (2) the combined HapMap3 and MEGA variant set.

### Genomic enrichment analysis

We assessed enrichment of breast cancer heritability in functional genomic annotations using stratified LD score regression (S-LDSC) with the baseline-LD v2.2 model^33–35^, which includes 73 binary annotations spanning regulatory, coding, and evolutionary features compiled from ENCODE, Roadmap Epigenomics, and comparative genomics resources. Analyses were restricted to HapMap3 variants with MAF > 0.05. For EAS and EUR, we used precomputed LD scores with baseline-LD annotations, and for AFR and H/L we computed ancestry-specific LD scores while using the same annotation definitions. Enrichment was defined as the proportion of heritability explained by variants in an annotation divided by the proportion of variants overlapping the annotation. Additional details are provided in the **Supplementary Methods**.

### Cell-type specific associations analysis using scDRS+

To connect genetic risk to cellular contexts, we applied scDRS+^36,37^, an extension of scDRS^36^ with improved accuracy through single-cell imputation and modeling of related cell populations, to GWAS summary statistics from each ancestry and the Tabula Sapiens single-cell RNA-seq atlas^38^. We restricted to fluorescence-activated cell sorting (FACS) cells to improve data quality. For each ancestry, we prioritized the top 1,000 genes ranked by MAGMA (Multi-marker Analysis of GenoMic Annotation)^39^ and computed cell-level disease relevance scores, followed by cell-type enrichment testing via aggregation of cell-level scores. We identified marginally associated cells using an FDR threshold of 0.1 and then applied joint fine-mapping to prioritize associated cells shared across ancestries. Details on atlas filtering, variant sets, concordance analyses across ancestries, and statistical testing are provided in the **Supplementary Methods**.

## RESULTS

### Logit-scale SNP-based heritability

We estimated logit-scale SNP-based heritability (*h*^2^) within each ancestry group using LDSC (**Table 1**). The estimates were *h*^2^ =0.614 (SE = 0.095) in AFR, *h*^2^ =0.466 (SE = 0.066) in EAS, EUR *h*^2^ =0.501 (SE = 0.050) and H/L *h*^2^ =0.588 (SE = 0.360). Despite modest differences in point estimates, there was no evidence of heterogeneity in SNP-based heritability across ancestry groups (Cochran’s Q = 1.72, p = 0.63; **Figure S2**).

Sensitivity analyses varying the variant set (HapMap3 vs HapMap3 plus MEGA) and MAF threshold (MAF > 0.01 vs MAF > 0.05) produced consistent estimates in EAS and EUR (**Table S2**). In AFR, estimates varied with the LD reference panel and were moderately higher at MAF > 0.01 than at MAF > 0.05 when the primary AABCG reference was used; we therefore used the AABCG-based estimate as primary and report 1000 Genomes AFR results as a sensitivity analysis. H/L estimates were imprecise and varied across MAF thresholds and variant sets; given the smaller sample, we used MAF > 0.05 to focus on more reliably estimated common variants and HapMap3 plus MEGA to retain broader common-variant coverage.

### Polygenicity and projected PRS performance

Using GENESIS, the mixture model converged for AFR, EAS, and EUR but not for H/L. The available H/L summary statistics and LD inputs did not support stable estimation under the three-component mixture model; therefore, polygenicity and projected CT-PRS results are reported only for AFR, EAS, and EUR.

After GENESIS preprocessing excluded the MHC region, SNP-level extreme-effect outliers with Z^2^ > 80, and variants with low effective sample sizes, the fitted effect-size distributions showed no evidence of inflation across the ancestry groups in which the model converged (**Figure S3**). The model-implied number of non-null susceptibility SNPs among the analyzed HapMap3 variants was 8,308 (SE = 2,751) in AFR, 4,446 (SE = 3,100) in EAS, and 5,235 (SE = 1,191) in EUR (**Table S3**). Differences were not statistically significant across ancestries (Cochran’s Q = 1.20, p = 0.55), indicating that observed variation is consistent with sampling variability and differences in power or LD structure.

We next used GENESIS to project CT-PRS performance as a function of increasing GWAS sample size (**Figure 1, Table S4**). At a total sample size of *N* = 200,000, comprising 100,000 cases and 100,000 controls, the projected proportions of SNP-based heritability captured by CT-PRS were 26.2% in AFR, 39.4% in EAS, and 38.6% in EUR. The corresponding projected AUCs were 0.611, 0.634, and 0.621, respectively (**Figure 1, Table S4**). Assuming the same 1:1 case-control ratio, projected AUCs at *N* = 500,000 were 0.660 in AFR, 0.664 in EAS, and 0.651 in EUR; at *N* = 1,000,000, they were 0.681, 0.675, and 0.666, respectively. Thus, doubling the sample size from 500,000 to 1,000,000 produced only modest gains, and the projected AUCs remained below their asymptotic values of approximately 0.71 in AFR and EAS and 0.69 in EUR (**Table S3**).

**Figure 1.** Model-based projections of genetic variance explained and predictive performance of clumping-and-thresholding polygenic risk scores (CT-PRS) under the GENESIS framework across hypothetical GWAS sample sizes. A) Estimated proportion of GWAS heritability explained by CT-PRS as a function of total GWAS sample size. The proportion was calculated as the variance explained by CT-PRS divided by the logit-scale heritability of common variants. B) Projected area under the receiver operating characteristic curve (AUC) for CT-PRS at different sample sizes. Detailed projections are provided in Table S4. The red dashed line indicates a total sample size of 200,000 individuals (100,000 cases and 100,000 controls). Abbreviations: AFR, African; EAS, East Asian; EUR, European.

To assess whether the model-based projections were consistent with published empirical performance, we compared the projected CT-PRS AUCs with observed AUCs from ancestry-specific PRS studies at the corresponding reported training sample sizes (**Table S5**)^19,20,40^. In AFR, GENESIS projected an AUC of 0.546 for 12,174 training cases and 13,160 training controls, compared with an empirical AUC of 0.560 for a single-ancestry CT-PRS^20^. In EAS, the projected AUC was 0.586 for 22,013 cases and 22,114 controls, compared with empirical AUCs of 0.589 in a case-control validation dataset and 0.600 in prospective cohorts for a CT-PRS^40^. In EUR, the projected CT-PRS AUC was 0.611 for 84,668 cases and 67,515 controls, compared with an empirical AUC of 0.630 for PRS313, which extended a CT-PRS through stepwise selection^19^. Across these comparisons, the empirical AUCs were within 0.003-0.019 of the corresponding GENESIS projections. This close agreement supports the GENESIS curves as realistic reference trajectories for common-variant CT-PRS prediction, while recognizing differences in evaluation design and PRS construction.

### Cross-ancestry genetic correlations of breast cancer

We estimated cross-ancestry genetic correlations using Popcorn (**Figure 2, Table S6**). Using HapMap3 variants, correlations were highest between EAS and EUR (*ρ* = 0.79, SE = 0.08) and between EUR and H/L (*ρ* = 0.68, SE = 0.21). Correlations involving AFR were lower, ranging from *ρ* = 0.26 (SE = 0.24) between AFR and H/L to *ρ* = 0.42 (SE = 0.14) between AFR and EUR. Results were similar when extending the variant set to HapMap3 plus MEGA (**Figure 2B, Table S6**).

**Figure 2.** Heatmaps of cross-sample genetic correlations for breast cancer estimated by Popcorn using A) HapMap3 variants and B) HapMap3 and Multi-Ethnic Genotyping Array variants combined. Abbreviations: AFR, African; EAS, East Asian; EUR, European; H/L, Hispanic/Latina.

### Genomic enrichment of breast cancer heritability

S-LDSC identified enrichment of breast cancer heritability in regulatory annotations, with the strongest signals in EUR (**Table S7**). In EUR, ancient promoter regions were highly enriched (32.42-fold, p = 1.98×10^-4^), along with transcription factor binding sites (5.79-fold, p = 9.16×10^-5^) and H3K4me3-marked promoters (4.90-fold, p = 8.56×10^□5^). Additional significant enrichment was observed for super-enhancers (3.01-fold, p = 3.73×10^□11^) and enhancer-associated marks H3K4me1 (2.52-fold, p = 1.28×10^□7^) and H3K27ac (2.16-fold, p = 2.25×10^□10^). In EAS, super-enhancers were significantly enriched after Bonferroni correction (2.73-fold, p = 2.60×10^□7^), and in AFR, H3K27ac showed significant enrichment (2.15-fold, p = 2.59×10^□4^). No annotation reached statistical significance in H/L, consistent with limited power. Across annotations that were significant in at least one ancestry group, enrichment patterns were qualitatively concordant across ancestries (**Figure S4**), and heterogeneity tests did not identify significant cross-ancestry differences after multiple-testing correction.

### Cell-type specific associations with breast cancer risk

We applied scDRS+ to integrate ancestry-specific GWAS results with the Tabula Sapiens single-cell atlas and identify cell types enriched for expression of GWAS-prioritized genes (**Figure 3, Tables S8-S9**). Because closely related cell populations can yield correlated signals, we report both fine-mapped results and marginal cell-type enrichments.

**Figure 3.** scDRS+ results using Tabula Sapiens. One asterisk indicates FDR < 0.1; two asterisks indicate FDR < 0.05. A) Fine-mapped associations, showing FDR values for cell types in which at least one ancestry had FDR < 0.05; all displayed associations were also marginally significant. B) Marginal associations, showing FDR values for cell types associated across all ancestries. C) UMAP of the cell types shown in panel A. D) UMAPs of scDRS+ scores across ancestries (AFR, EAS, EUR, H/L) for the same cell types as in panel C. Red indicates greater disease relevance.

Across ancestries, we observed consistent associations in innate immune cell types, including classical monocytes and neutrophils (**Figure 3A, B**), supporting involvement of myeloid-related programs in genetic susceptibility. In addition, scDRS+ highlighted cell populations annotated outside mammary tissue, including lacrimal gland functional unit cells and skeletal muscle satellite stem cells (**Figure 3A**). These associations are best interpreted as atlas-dependent, hypothesis-generating signals reflecting the multi-tissue nature of the Tabula Sapiens reference rather than direct etiologic tissues. Consistent with this interpretation, mammary luminal epithelial cells and breast fibroblasts showed marginal associations in multiple ancestries, although they were not prioritized by fine-mapping (**Figure 3B, Table S9**).

To evaluate cross-ancestry concordance, we compared marginal scDRS+ scores across ancestry pairs within key associated cell types and observed positive correlations for classical monocytes, neutrophils, and skeletal muscle satellite stem cells (**Figure S5**). UMAP visualizations similarly demonstrated overlapping disease-relevant cell clusters across ancestries (**Figure 3C, D**).

## DISCUSSION

We present a systematic cross-ancestry evaluation of breast cancer genetic architecture using GWAS summary statistics from AFR, EAS, EUR, and H/L samples. Across multiple complementary analyses, we find evidence for substantial sharing of common-variant architecture across ancestries, with broadly comparable logit-scale SNP-based heritability and polygenicity estimates, consistent regulatory enrichment signals, and convergent cellular contexts highlighted by single-cell integration. Although point estimates varied across samples, formal heterogeneity tests did not identify significant cross-ancestry differences, suggesting that much of the observed variation may be attributable to differences in power, LD structure, and reference panel choice rather than large differences in underlying biology.

Breast cancer exhibited moderate logit-scale SNP-based heritability across all groups, with the highest point estimate in AFR. Estimates were stable in EAS and EUR across sensitivity analyses, but were more sensitive to the LD reference panel in AFR, where using sample-matched LD from AABCG controls increased heritability relative to using 1000 Genomes AFR (**Table S2**). AFR estimates were also moderately sensitive to the MAF threshold, adding practical uncertainty to direct cross-ancestry comparisons involving AFR. We selected AABCG controls as the primary AFR LD reference because they more closely matched the predominantly U.S.-based African American GWAS than the smaller 1000 Genomes AFR panel. Despite this sensitivity, we found no evidence of heterogeneity in SNP-based heritability across ancestry groups. This highlights the importance of appropriate ancestry-specific LD modeling in admixed populations^41–43^. Despite modest variation in point estimates, we found no evidence of heterogeneity in SNP-based heritability across samples (p = 0.63). Our estimates are broadly consistent with prior reports based on subsets of these data. For example, Michailidou et al. (2017)^7^, estimated that common variants explain about 41% of the familial relative risk in large European-ancestry analyses, corresponding to a logit-scale heritability of approximately 0.568, similar to our EUR estimate (0.501). Likewise, our AFR estimate of 0.614 is comparable to the logit-scale heritability (referred to as frailty-scale) of 0.667 reported in the recent African ancestry sample breast cancer GWAS^12^, which used the same summary statistics but a different LD reference panel. While heritability has been estimated in prior studies, these have largely focused on individual ancestry groups and used varying methodologies; to our knowledge, our study provides the first systematic, cross-ancestry comparison of logit-scale SNP-based heritability using harmonized models. We focused on the logit scale, as it provides an interpretable and robust estimate of genetic variance explained by GWAS variants and connects directly to familial risk^44^, without requiring assumptions about disease prevalence.

Following GENESIS guidance, analyses in all ancestry groups were restricted to HapMap3 variants with MAF > 0.05. This harmonized restriction supports consistent mixture-model fitting across ancestries but limits inference to common, well-tagged variants and may exclude lower-frequency or population-specific contributions, particularly in AFR. Accordingly, the model-implied polygenicity and projected CT-PRS performance describe the common-variant architecture represented by the analyzed HapMap3 variants rather than the complete allelic architecture of breast cancer risk or the performance potentially achievable with broader variant coverage.

With this common-variant scope, GENESIS suggested substantial polygenicity across ancestry groups, with model-implied numbers of non-null susceptibility SNPs ranging from 4,446 in EAS to 8,308 in AFR, though differences were not statistically significant (p = 0.55). These quantities represent susceptibility signals captured by the analyzed HapMap3 variant set rather than the total number of causal variants across the allele-frequency spectrum. Translating this architecture into sample-size-dependent CT-PRS trajectories showed that projected AUC increased with GWAS sample size but approached the asymptote slowly. Although the estimated asymptotic AUC in AFR was comparable to those in EAS and EUR, the AFR curve rose more slowly at currently achieved sample sizes, consistent with empirically observed lower PRS performance in AFR samples at current GWAS scales^20,45^. These projections help distinguish limitations related to discovery sample size from those related to the common-variant architecture represented by the model. Improving PRS performance in underrepresented populations will require larger ancestry-diverse GWAS together with improved variant coverage, imputation panels, and ancestry-matched LD resources^46^.

Published empirical AUCs were close to the GENESIS projections at comparable reported training sample sizes, supporting the curves as reasonable reference trajectories for common-variant CT-PRS prediction rather than substitutes for external validation (**Table S5**). The projections are specific to CT-PRS and should not be interpreted as upper bounds for other prediction methods. More flexible approaches, including LD-aware shrinkage, multi-ancestry integration, subtype-aware modeling, and penalized regression, have achieved higher external discrimination^18–20^.

Genetic correlations were strongest between EUR and EAS (*ρ* = 0.79), and lower in comparisons involving AFR, consistent with ancestry differences in LD and allele-frequency patterns that affect variant tagging and reflect broad population divergence^47^. The moderately strong correlation between EUR and H/L (*ρ*= 0.68) likely reflects European ancestry contributions in many H/L cohorts. Thus, these correlations should be interpreted as average effect-size sharing within heterogeneous groups. More broadly, both AFR and H/L summary statistics represent genetically heterogeneous groups. The AFR data largely reflect African American cohorts with predominantly West African and European admixture; performance and correlation estimates from these data should not be extrapolated to populations of South, East, or Central African ancestry, where LD patterns and causal variant frequencies may differ substantially. Similarly, the H/L aggregate estimate likely masks heterogeneity across individuals and cohorts with varying proportions of Indigenous American, African, and European ancestry. Future analyses incorporating local ancestry or ancestry-stratified GWAS will provide finer resolution of these correlation structures. Because correlations involving H/L were restricted to variants with MAF > 0.05 in the H/L sample, they primarily reflect genetic sharing among variants common in H/L. This threshold yields larger minor-allele counts and more precise effect estimates in the relatively small H/L GWAS, but excludes contributions from variants with H/L MAF between 0.01 and 0.05.

Stratified heritability analyses indicated broadly consistent enrichment of regulatory annotations across ancestries, including promoter- and enhancer-related elements and active chromatin marks such as H3K27ac and H3K4me1. Although enrichment magnitudes differed, heterogeneity tests did not identify significant cross-ancestry differences among annotations reaching significance in at least one group, and no annotation reached significance in H/L, consistent with limited power.

Integrating GWAS with single-cell transcriptomic data using scDRS+^37^, we observed concordant signals in classical monocytes and neutrophils, as well as in lacrimal gland functional unit cells and skeletal muscle satellite stem cells. The myeloid signals align with prior evidence implicating monocytes and neutrophils in tumor inflammation and progression^48–53^. Signals in cell types annotated outside mammary tissue should be viewed as atlas-dependent hypotheses rather than evidence that these tissues are etiologic; their interpretation may change as more comprehensive and breast-specific single-cell atlases become available. Marginal associations in mammary luminal epithelial cells and breast fibroblasts provide complementary evidence for epithelial and stromal contexts. The concordance across ancestries supports shared cellular programs, but does not establish that the implicated cell types mediate genetic risk.

This is the largest cross-ancestry investigation of breast cancer architecture; however, several limitations exist. First, the study did not train or externally validate a new PRS. GENESIS therefore provides model-based expectations conditional on the specified architecture and analyzed variant set, not empirical PRS performance in independent data. Second, we did not evaluate clinical-utility measures, such as the proportion of individuals exceeding a risk threshold for enhanced screening. Translating projected AUC to absolute risk would require an externally validated score, target-population calibration, age-specific baseline incidence, and a prespecified clinical threshold. Third, statistical power remained limited in non-European samples, particularly H/L, where GENESIS did not converge and enrichment analyses did not reach significance. The H/L non-convergence indicates that the available summary statistics and LD inputs were insufficient for stable estimation under this model and should not be interpreted as evidence of a fundamentally distinct genetic architecture.

These limitations highlight the value of expanding breast cancer GWAS in diverse populations. <u>The Confluence Project</u>, which aims to assemble approximately 300,000 cases and 300,000 controls through collaboration across major consortia, should enable more precise inference and more generalizable genetic risk models. In summary, our study refines the understanding of breast cancer genetics across populations and highlights the importance of multi-ancestry studies for etiologic insight and polygenic risk prediction.

## ACKNOWLEDGEMENTS

The analysis utilized DNANexus and the high-performance computation Biowulf cluster at the National Institutes of Health (NIH), USA. We would like to thank the following members of COLUMBUS Consortium:

University of California at Davis: Ana Estrada-Florez, Paul Lott, Guadalupe Polanco-Echeverry, Luis Carvajal-Carmona.

Colombia: Juan Manuel Acosta (Universidad del Tolima, Ibagué), Jennyfer Benavides (Universidad del Tolima, Ibagué), Mabel Bohorquez (Universidad del Tolima, Ibagué), Jenny Carmona (Dinámica IPS, Medellín), Ángel Criollo (Universidad del Tolima, Ibagué), Magdalena Echeverry (Universidad del Tolima, Ibagué), Ana Estrada-Florez (Universidad del Tolima, Ibagué), Gilbert Mateus (Hospital Federico Lleras Acosta, Ibagué), Raúl Murillo (Pontificia Universidad Javeriana, Bogotá,), Justo Ramirez (Hospital Hernando Moncaleano Perdomo, Neiva), Carolina Sanabria (Instituto Nacional de Cancerología, Bogotá), Yesid Sánchez (Universidad del Tolima, Ibagué), Martha Lucia Serrano (Instituto Nacional de Cancerología, Bogotá), John Jairo Suarez (Universidad del Tolima, Ibagué), Alejandro Vélez (Dinámica IPS, Medellín, Colombia, Hospital Pablo Tobón Uribe, Medellín).

Mexico: Javier Torres (Mexican Social Security Hospital).

The funder had no role in the design of the study; the collection, analysis, or interpretation of the data; or the writing of the manuscript and decision to submit it for publication.

## AUTHOR CONTRIBUTIONS

T.U.A and H.Z. conceived the project. J.L.L. and J.Williams conducted the heritability analyses. J.Williams, Q.H. and L.Y. conducted the polygenicity analyses. O.J. conducted the cross-sample genetic correlation analyses. J.Williams and M.Z. conducted the enrichment analyses. A.T. conducted scDRS+ analyses. J.W. computed the LD-scores for EAS, EUR and H/L. G.J. computed the LD-scores and MAGMA analyses on AABCG controls. Statistical analyses were conducted under the guidance of K.M., Q.Z., M.J.Z., D.D., T.U.A., and H.Z.; J.L.L., M.J.Z., J. Williams, O.J., A.T., T.U.A. and H.Z. wrote the first draft. J.T.B., L.F., W.Z., S.J.C., N.C., P.K., M.G.C., M.J.Z., T.U.A and H.Z. reviewed and edited the draft. All co-authors reviewed and approved the final version of the manuscript.

## DATA AVAILABILITY

Harmonized, quality-controlled breast cancer GWAS summary statistics for all four ancestry groups, restricted to HapMap3 and MEGA variants are available through Harvard Dataverse (https://dataverse.harvard.edu/dataset.xhtml?persistentId=doi:10.7910/DVN/N1WIFR). AFR GWAS summary statistics are available at GWAS Catalog (<u>GCST90296719</u>)

BCAC EAS GWAS summary statistics are available at: https://www.ccge.medschl.cam.ac.uk/breast-cancer-association-consortium-bcac/data-data-access/summary-results/gwas-summary-results

Biobank Japan GWAS summary statistics are available at: https://pheweb.jp/pheno/BrC

BCAC EUR GWAS summary statistics are available at: https://www.ccge.medschl.cam.ac.uk/breast-cancer-association-consortium-bcac/data-data-access/summary-results/gwas-summary-associations

The H/L GWAS summary statistics are available by contacting Dr. Elad Ziv to gain access.

## CODE AVAILABILITY

All data analysis code is available at: https://github.com/confluence-breast-cancer-consortia/pre-confluence-analysis/tree/main

LDSC and stratified LDSC are available at: https://github.com/bulik/ldsc

GENESIS is available at: https://github.com/yandorazhang/GENESIS

Popcorn is available at: https://github.com/brielin/Popcorn

scDRS is available at: https://github.com/martinjzhang/scDRS

