## Supplementary Figures for "Genetic and Cellular Architecture of Breast Cancer Risk Across Ancestries"

### Supplementary Material

**Figure S1. Workflow for processing genome-wide association study (GWAS) summary statistics.** Note: Minor allele frequency, MAF; Multi-Ethnic Genotyping Array, MEGA

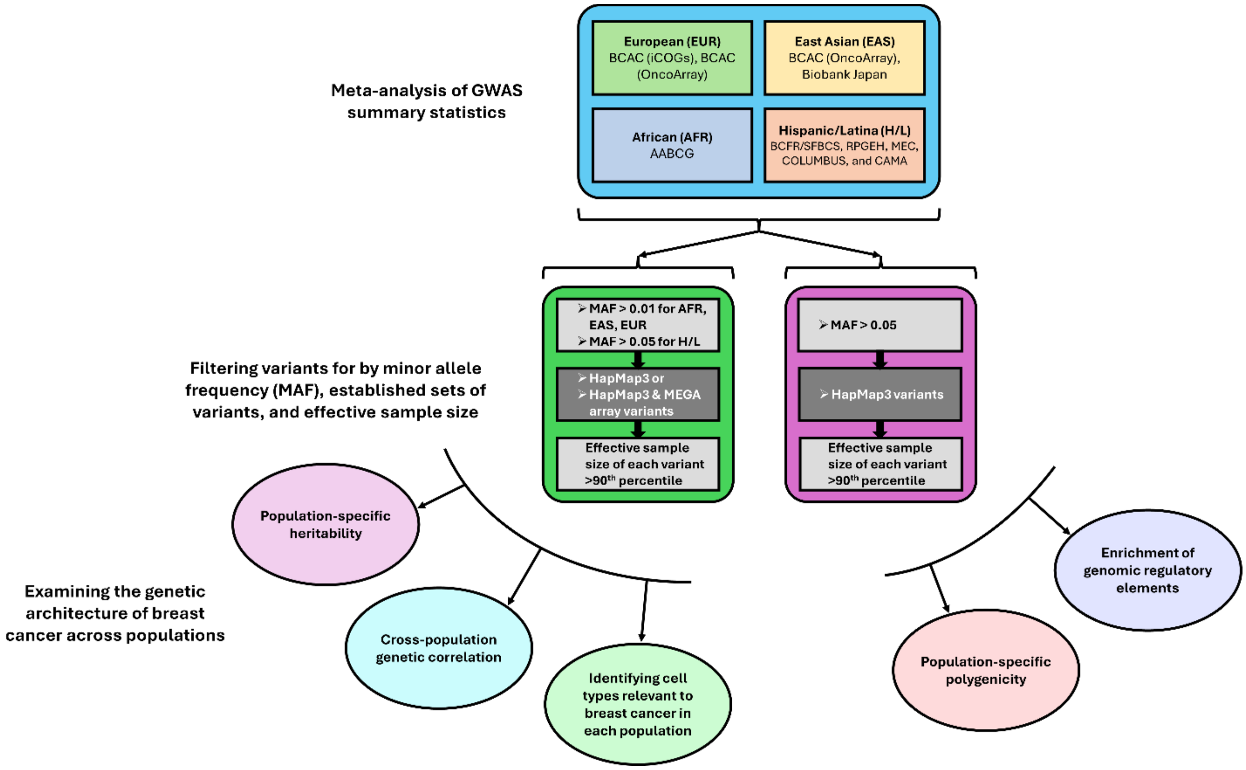

**Figure S2. Forest plot of sample-specific estimates of the logit-scale SNP-based heritability**. The logit-scale heritability (also known as frailty-scale heritability) is defined as $\sigma^2_{GWAS} = Var(\sum_{m=1}^{M} \beta_m G_m)$, where $G_m$ is the standardized genotype for the mth SNP, $\beta_m$ is the true log odds ratio for the mth SNP and M is the total number of causal SNPs among the GWAS variants. AFR, African; EAS, East Asian; EUR, European; H/L, Hispanic/Latina. Error bars represent ±1 standard error. A heterogeneity test across ancestry-specific heritability estimates yielded Cochran's Q = 1.72 (p = 0.63), indicating no significant evidence of heterogeneity.

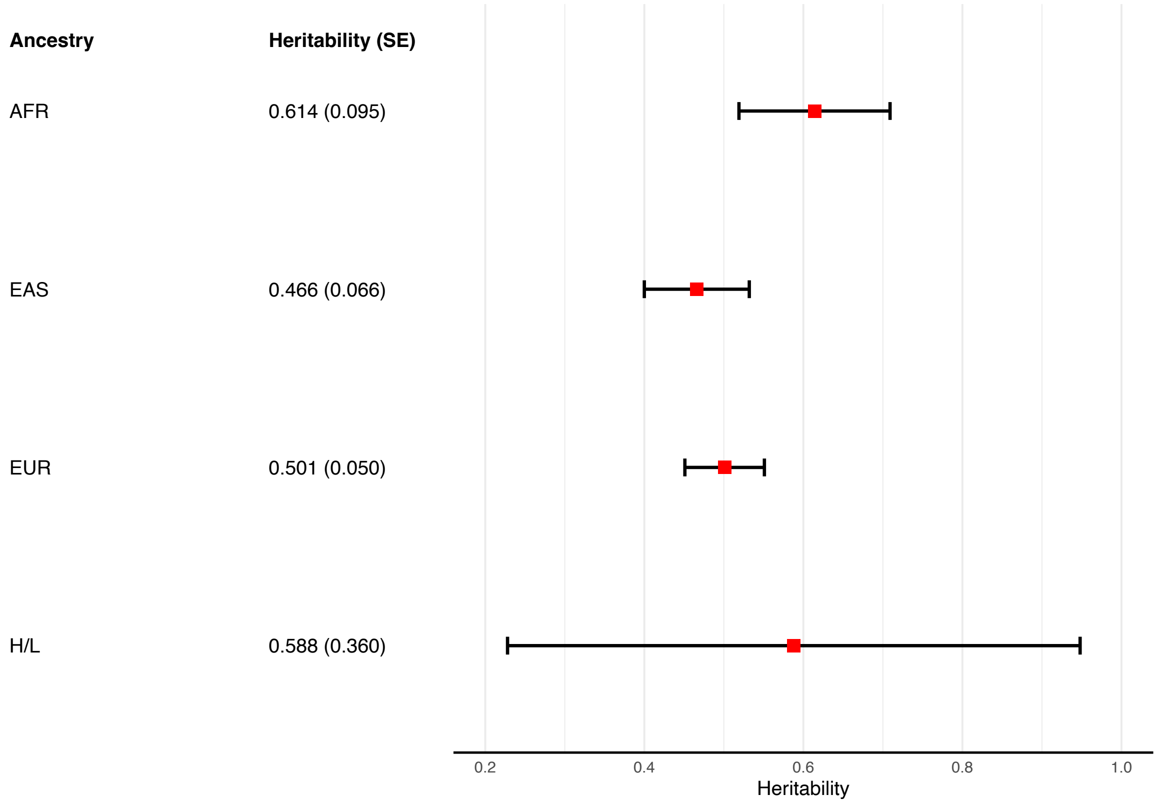

**Figure S3. Q-Q plots comparing the observed association statistic distributions and those expected under the three-component model fit by GENESIS**. A) African B) East Asian C) European

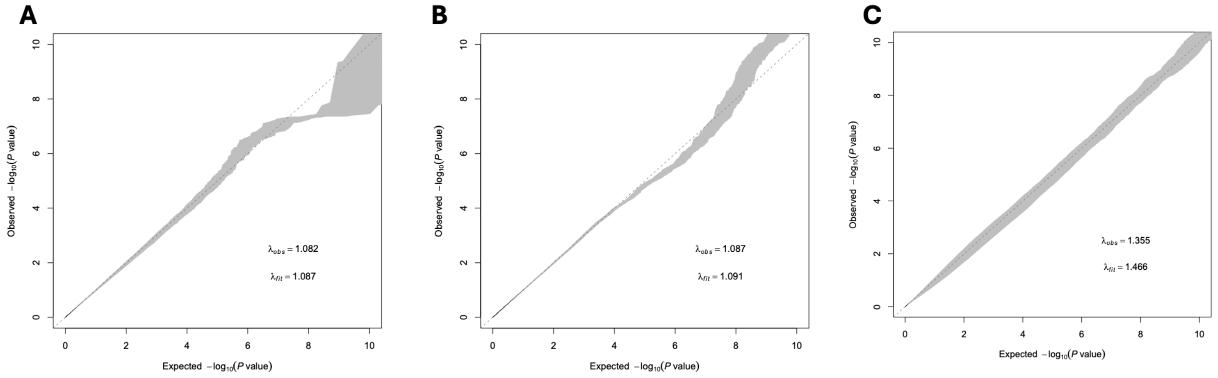

**Figure S4. Enrichment analysis results for significantly enriched annotations.** Error bars represent Jackknife standard errors around the estimates of enrichment. The chi-square test was used to assess heterogeneity between ancestries. (Transcription factor for binding site (TFBS), $p = 0.82$; H3K4me3, $p = 0.483$; Super Enhancer, $p = 0.194$; H3K4me1, $p = 0.032$; H3K27ac, $p = 0.945$)

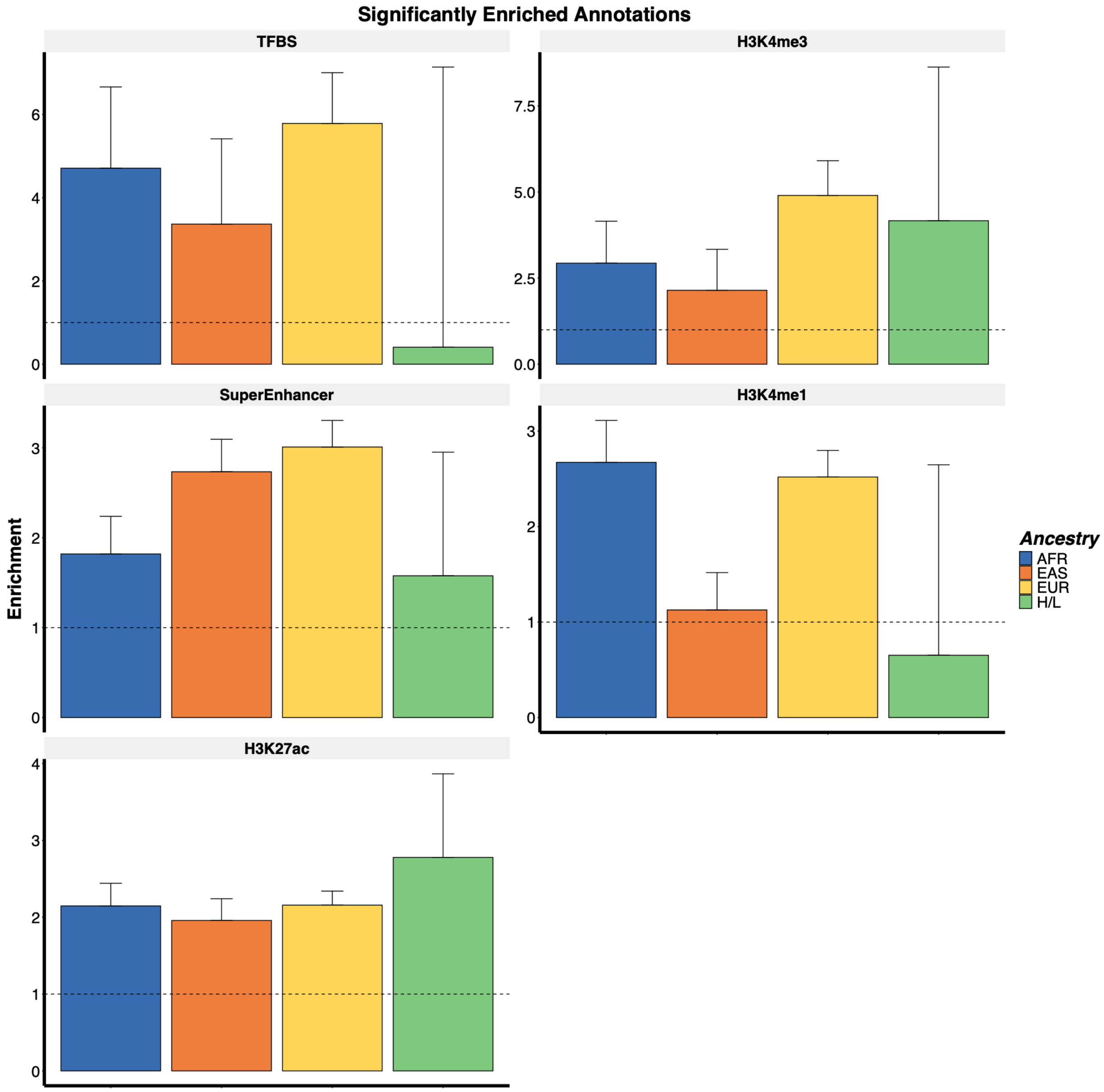

**Figure S5. Correlation of scDRS+ scores between ancestries in A) neutrophils, B) classical monocytes, C) skeletal muscle satellite stem cells, and D) macrophages**. Significance of correlation between ancestries assessed through empirical p-values in E) neutrophils, F) classical monocytes, G) skeletal muscle satellite stem cells, and H) macrophages.

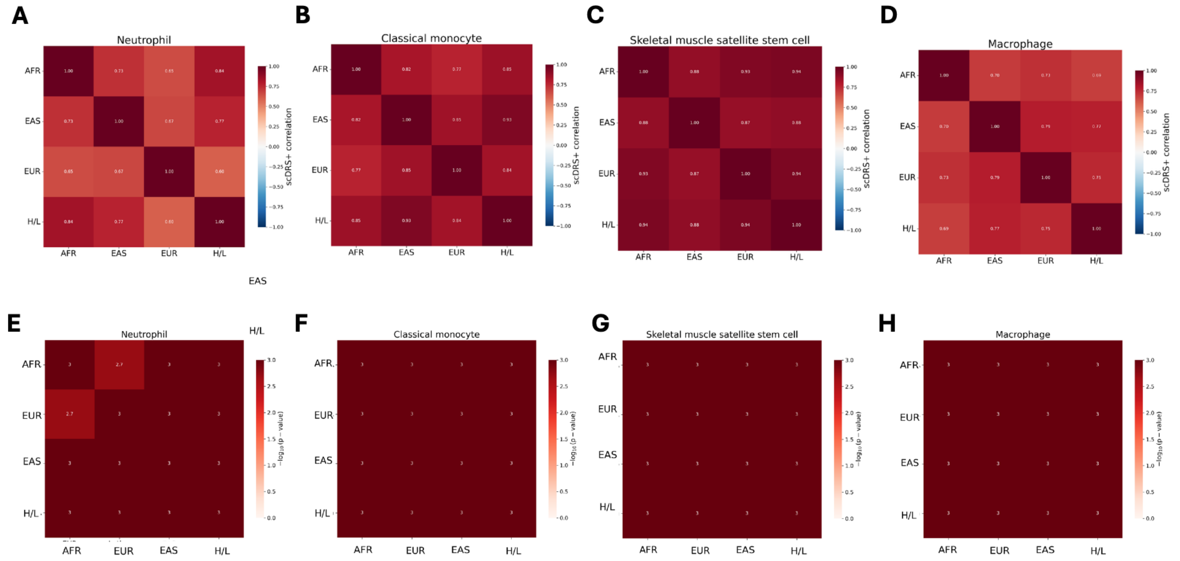
